# Selected miRNAs in urinary extracellular vesicles show promise for non-invasive diagnostics of diabetic kidney disease

**DOI:** 10.1101/2024.09.12.24312889

**Authors:** Karina Barreiro, Jenni Karttunen, Erkka Valo, Essi Viippola, Ileana Quintero, Annemari Käräjämäki, Antti Rannikko, Harry Holthöfer, Andrea Ganna, Niina Sandholm, Lena M. Thorn, Per-Henrik Groop, Tiinamaija Tuomi, Om Prakash Dwivedi, Maija Puhka

**Author notes:** shared first author (Karina Barreiro, Jenni Karttunen). **Correspondence**: Maija Puhka, Associate Professor, PhD, Senior Scientist, Head of HiPREP core in FIMM Technology Centre, Institute for Molecular Medicine Finland FIMM, HiLIFE, University of Helsinki, Biomedicum 2U, Tukholmankatu 8, 00290 Helsinki, Finland.

## Abstract

Diabetic kidney disease is a growing health burden that lacks specific early non-invasive diagnostic procedures. To approach a solution for this clinical need, we sequenced microRNAs of urinary extracellular vesicles and performed biomarker discovery by small RNA sequencing in a type 1 diabetes cohort including males with and without albuminuria. The results were replicated by sequencing or qPCR in two independent cohorts and four previously published datasets including type 1 and 2 diabetes as well as both sexes. Non-diabetic and prostate cancer cohorts were used as additional controls and miRNAs changed due to preanalytical urine collection variables were excluded. Using these data, we additionally validated previously identified reference candidate miRNAs. Correlations with clinical parameters, receiver operating characteristic analysis, targeted mRNAs and pathways including integrated single cell data, and targeted circulating proteins from type 1 and 2 diabetes cohorts were analyzed. We pinpointed 6 stable microRNAs, 11 differentially expressed microRNAs, 9 target proteins and 16 DKD-associated pathways in individuals with diabetic kidney disease. Replication showed that the differentially expressed miRNAs in DKD were partly shared between diabetes subtypes and sexes with overall strongest evidence for miR-192-5p, miR-146a-5p, miR-486-5p, and miR-574-5p. Combination of these miRNAs with clinical variables showed potential to classify individuals with the fastest kidney function decline (sensitivity 0.75-1.00 and specificity 0.83-1.00) even in the normoalbuminuria group, thus holding the potential as early diagnostic markers. Altogether, the candidate microRNAs show specificity for diabetic kidney disease, identify declining kidney function, and target key kidney cell types, mRNAs, proteins, and pathogenic mechanisms.

**Lay summary:** Diabetic kidney disease (DKD) damages the kidneys severely. DKD progression differs between diabetes subtypes and sexes and currently the diagnosis comes too late when the kidney is already damaged. Thus, clinical care needs better and earlier diagnostic markers. Here we studied the microRNAs in urinary extracellular vesicles, small particles secreted actively by the kidney cells. We found 11 candidate microRNA markers for DKD. The discovery was made in males with type 1 diabetes, but some of the microRNAs were confirmed across type 1 and 2 diabetes, as well as female or combined sex populations. Altogether we studied over 250 samples. Analysis of miRNA’s regulatory pathways and correlations with clinical measurements in individuals with DKD suggest that some miRNAs may hold the potential as early diagnostic markers. Our findings could thus impact clinical practice by providing early and specific DKD diagnostic tools allowing effective care at an early DKD stage.

## Introduction

Diabetes is a growing health burden that affects close to 10% of the world population.^1^ Particularly problematic is that devastating diabetic complications cannot be sufficiently prevented nor cured. Diabetic kidney disease (DKD) is a common microvascular complication of ∼30% of the individuals with type 1 diabetes (T1D)^2,3^ and ∼40% of those with type 2 diabetes (T2D)^3^. The factors leading to DKD include hyperglycemia, oxidative stress, chronic inflammation, obesity, insulin resistance, hypertension, but also genetic factors^4–6^. Of note, the DKD susceptibility and disease course differs between the sexes^7,8^ and diabetes subtypes^9^. DKD manifests as progressive albuminuria and a relentless decline in the kidney function measured by estimated glomerular filtration rate (eGFR).^10^ As the diagnosis currently relies on the rather late and somewhat unspecific biomarkers albuminuria and eGFR, the kidneys are already to some extent damaged at the time of diagnosis.^10^ Kidney biopsies could provide an accurate diagnosis, but a kidney biopsy is an expensive and invasive procedure with a risk of complications.^11,12^ Novel, early and specific diagnostics would enable early interventions to protect the kidney - combatting the progression of DKD to kidney failure.

Urinary extracellular vesicles (uEV) offer a potential non-invasive diagnostic solution. The uEV are secreted nanovesicles that carry a rich RNA cargo reflecting the pathophysiological state of the originating genitourinary cells and tissues^13^. Sex affects the uEV secretion and cargo: males produce more kidney-derived uEV and generally uEV, with partly different nucleic acids than females^14–17^. Our previous DKD studies indicated that the uEV represent a useful source of kidney miRNAs and mRNAs.^15,18^ In both sexes, uEV carry RNAs showing correlations with e.g., eGFR, albuminuria, and CKD stage.^15,18,19^ However, several technical challenges and unanswered questions in uEV-based DKD research still remain. Generally, the uEV studies have been small and have applied variable preanalytical, analytical, and clinical study designs.^13,20^ DKD-linked uEV miRNAs have been studied more in T2D than in T1D. Furthermore, exploration of the candidate markers in multiple cohorts or control groups has been rare. Therefore, the biomarker candidates have varied even if some results appear initially replicable.^18^ Thus, our aim was to understand whether the uEV could provide miRNA biomarkers specifically for early or late DKD and for various diabetes subtypes, different sexes, or particular pathogenic pathways. With miRNA sequencing, we here discovered uEV miRNAs linked with DKD, declining kidney function and key pathogenic mechanisms.

## Materials and methods

### Urine samples from diabetes cohorts

Urine samples and clinical data for the discovery cohort and replication cohorts were collected in the nationwide prospective Finnish Diabetic Nephropathy (FinnDiane) Study and the Diabetes Registry in Vaasa (DIREVA study) as described^15^ (Table 1, Supplementary methods). Clinical data are presented for the T1D discovery and replication cohorts in Supplementary Table S1a and S1b, respectively, and for the T2D replication cohort in Table S3.^15^ The diagnosis, stratification criteria for the albuminuria group, and collection of clinical data has been described earlier^15^ and in the Supplementary methods.

**Table 1:** Cohorts profiled by miRNA sequencing in this study. Overnight urine collections (ON), protease inhibitors (PI), type 1 diabetes (T1D), type 2 diabetes (T2D), ultracentrifugation (UC), urinary extracellular vesicles (uEV). ** 24 h urine collections were not pre-cleared but overnight urine collections were pre-cleared.

| Cohort | Collection type | Storage Temp | PI * | Pre-Clearing | uEV isolation Method | DNA treatment | Disease and sex | n (Donors) | Previous omics profiling |
| --- | --- | --- | --- | --- | --- | --- | --- | --- | --- |
| Discovery cohort (FinnDiane & DIREVA) | 24 h/ON | -80 °C | yes | yes-no ** | UC | No | Non-diabetic and T1D. All males. | Non-diabetic=7<br>Normoalbuminuria=40<br>Microalbuminuria=15<br>Macroalbuminuria=19 | mRNA-seq<sup>15</sup> |
| Replication cohort T1D (FinnDiane) | 24 h | -80 °C | yes | yes | UC | Yes | T1D. All females. | Normoalbuminuria=23<br>Microalbuminuria=4<br>Macroalbuminuria=4 | mRNA-seq<sup>15</sup> |
| Replication cohort T2D (DIREVA) | ON | -80 °C | no | yes | UC | No | T2D. Females and males. | Microalbuminuria=14 (2 females)<br>Macroalbuminuria=9 (1 female) | mRNA-seq<sup>15</sup> |

### Extracellular vesicle isolation and analysis

The uEV were isolated by ultracentrifugation from 30 ml of urine in the T1D discovery and T2D replication cohorts^15^, and from 7.8 ml urine in the T1D replication cohort (Supplementary methods).

The uEV were analyzed using transmission electron microscopy (TEM) as described^21^ and with Immunoelectron microscopy (IEM), nanoparticle tracking analysis (NTA) and single particle interferometric reflectance imaging sensing (SP-IRIS) (Supplementary methods).

### RNA isolation, small RNA sequencing and quantitative PCR

Total uEV RNA was isolated using miRNEasy mini or micro kit (Qiagen, Hilden, Germany) and analyzed with Bioanalyzer 2100 (Agilent Technologies, Santa Clara, CA) (Supplementary methods). RNA sequencing libraries generation using Lexogen Small RNA-Seq library (Lexogen GmbH, Vienna, Austria) from 1 ng of RNA, sequencing data analysis, and qPCR with TaqMan assays (Table 2) has been described in the Supplementary methods.

**Table 2.**
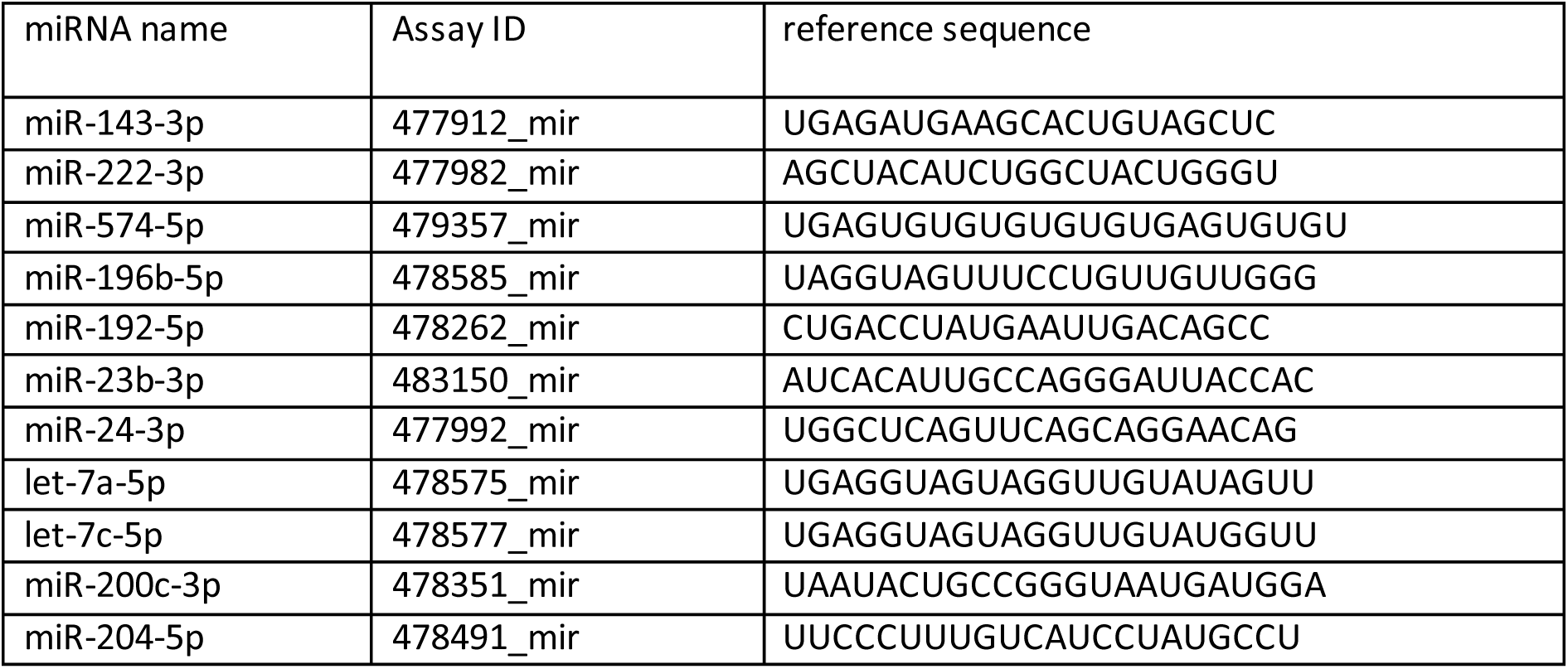
Details on TaqMan assays (Thermo Fisher Scientific) used for qPCR validation and normalization.

### Replication using published miRNA profiling datasets and receiver operating characteristic (ROC), pathway, and circulating protein analysis

To replicate our findings, we analyzed published datasets^22–27^ and our own data. We used IBM SPSS statistics V29 and CombiROC^28^ for ROC analysis, Ingenuity pathway analysis MicroRNA Target Filter (IPA, Qiagen) (accession date: 23.05.22) for pathway analysis and UK Biobank data^29^ for circulating plasma protein analysis. (Supplementary methods).

### Statistics

For sequencing data, exploratory and differential expression analyses were conducted using the R package DESeq2 v. 1.32.0^30^. Read count matrices were pre-filtered to exclude small RNAs with <10 reads in total across all samples. For heatmaps, principal component analysis (PCA), and violin plots data were transformed using variance stabilizing transformation (VST). MiRNAs that had a normalized mean counts >5, adjusted p-value (for multiple testing correction) <0.05, and absolute log2 fold change (FC) value >0.6 were considered differentially expressed (DE). Statistical testing for other analyses is described in the Supplementary methods.

## Results

In this study, we sequenced uEV miRNAs to identify biomarkers for DKD focusing on T1D and reference miRNAs not significantly affected by common preanalytical variables (Figure 1). The miRNAs were explored through various correlation analysis, ROC plots and target mRNA, circulating protein and pathway analysis incorporating single cell data.

**Figure 1.**
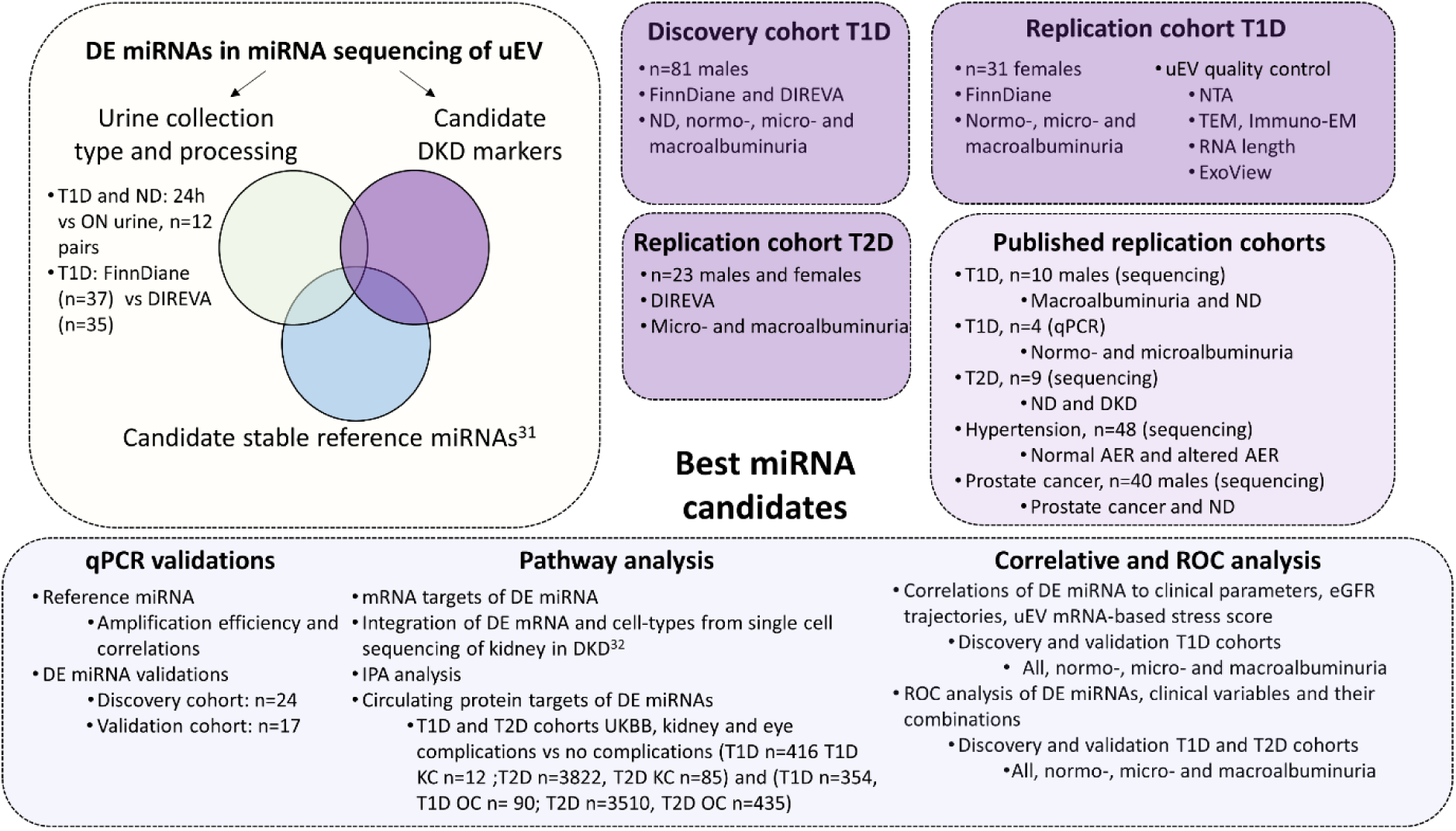
Study outline. The primary goal of the study was to discover urinary extracellular vesicle (uEV) miRNA markers for diabetic kidney disease (DKD) in individuals with T1D. The differentially expressed (DE) miRNAs were analyzed in the discovery and replication cohorts comprising different albuminuria groups and different timed urine collections. These data were cross-checked for stable expression of our previously identified reference miRNA candidates. ^31^ Candidate DKD marker and stable reference miRNAs were validated by qPCR. Markers were further explored in previously published uEV datasets and through integrative pathway, circulating protein target and ROC analysis. Expression levels were correlated with clinical measurements altered in DKD, eGFR trajectories reflecting long-term kidney function, and with a uEV-mRNA based “stress score”, which we have found to be elevated in DKD^15^. Albumin excretion ratio (AER), body-mass-index (BMI), estimated glomerular filtration rate (eGFR), hemoglobin A1c (HbA1c), kidney complications (KC), microalbuminuria (Micro), miRNA sequencing (miRNAseq), nanoparticle tracking analysis (NTA), non-diabetic (ND), normoalbuminuria (Normo), ophthalmic complications (OC), overnight (ON), receiver operating characteristic (ROC), transmission electron microscopy (TEM), type 1 diabetes (T1D), type 2 diabetes (T2D), United Kingdom Biobank (UKBB).

### uEV preparation and sequencing quality

The uEV were isolated and characterized as described^15^, except for the T1D replication cohort, where we used a protocol adopted for small urine volumes^31^ and uEV quality was ensured now. NTA showed 2.6 x 10^8^ – 3.1 x 10^9^ particles/ml of urine and a size distribution with a mean peak of 100 nm for all groups (Figure 2a). RNA presented typical uEV profiles: a main peak below ∼200 nt and occasionally, small ribosomal RNA peaks (Figure 2b). TEM revealed heterogenous uEV <500 nm in diameter (Figure 2b) and only few Tamm Horsfall Protein filaments. IEM and ExoView showed uEV positive for EV markers CD9, CD81 and CD63 (Figure 2c and Supplementary Figure S1). Exoview showed less CD81 than CD9 or CD63 positive particles, but all had equal sizes (Figure 2d, e). Only CD63 and CD9 co-positive particles were significantly decreased in the albuminuria samples (Figure 2f, g). Thus, the uEV preparation quality appeared fine as previously shown. Moreover, a good amount of raw sequencing reads and detected miRNAs were obtained for all cohorts (Supplementary results).

**Figure 2:**
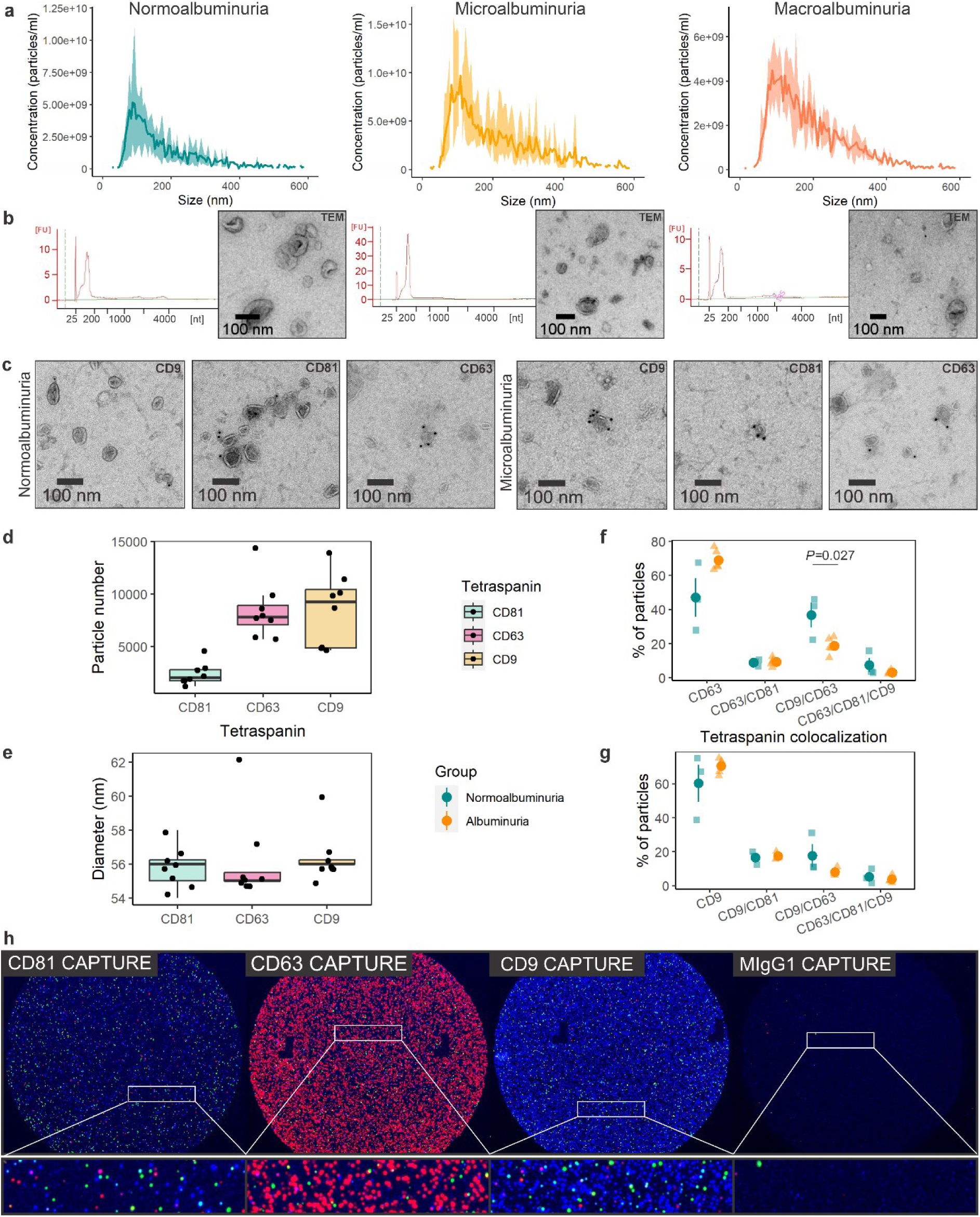
UEV characterization. (a) The particle size and concentration were measured using Particle Metrix Zetaview PMX-120 nanoparticle tracking analysis (n=3 per group). (b) Representative Agilent total RNA pico chip electropherograms used to assess uEV RNA quality and quantity and transmission electron microscopy micrographs close ups revealing particles with cup-shaped morphology. (c) Transmission electron microscopy micrograph close ups showing the immunodetection of CD9, CD81 and CD63. Wide-field micrographs for b and c are shown in Supplementary Figure S1. (d-h) Characterization of uEV with ExoView platform (n=8, normoalbuminuria n=3, albuminuria n=5). (d and e) Number and size of particles for CD81, CD63 and CD9 captured uEV. (f and g) Co-localization analysis for CD63 and CD9 captured uEV, respectively. The round point and vertical line represent the mean ± standard error of the mean. Due to the low number of CD81 particles, colocalization analysis is only shown for CD63 and CD9 captures. (h) Representative images of CD81, CD63, CD9, and IgG isotype control spots on the ExoView chips. Colored dots represent fluorescent detection of CD81 (green), CD63 (red), and CD9 (Blue) in particles captured in the sports. Urinary extracellular vesicles (uEV).

### Changes of the uEV miRNA profile in DKD

First, we compared different urine collections in the study (overnight vs 24 h and FinnDiane vs DIREVA, Supplementary results, Supplementary Figure S2 and Supplementary Tables S2a-c). In these preanalytical comparisons, we found 22 DE miRNAs, which were excluded when analyzing the DKD markers or reference miRNAs. Differential expression analysis for miRNAs in the T1D discovery cohort (Table 1) was started with the normo-vs macroalbuminuria group, revealing 11 DE miRNAs (Figure 3a, Supplementary Table S3b). Hierarchical clustering with the DE miRNAs showed clusters, although not a total separation of the groups (Figure 3b). The normalized counts of the DE miRNAs tended to increase from normo-to micro- and from micro-to macroalbuminuria (Figure 3c).

**Figure 3:**
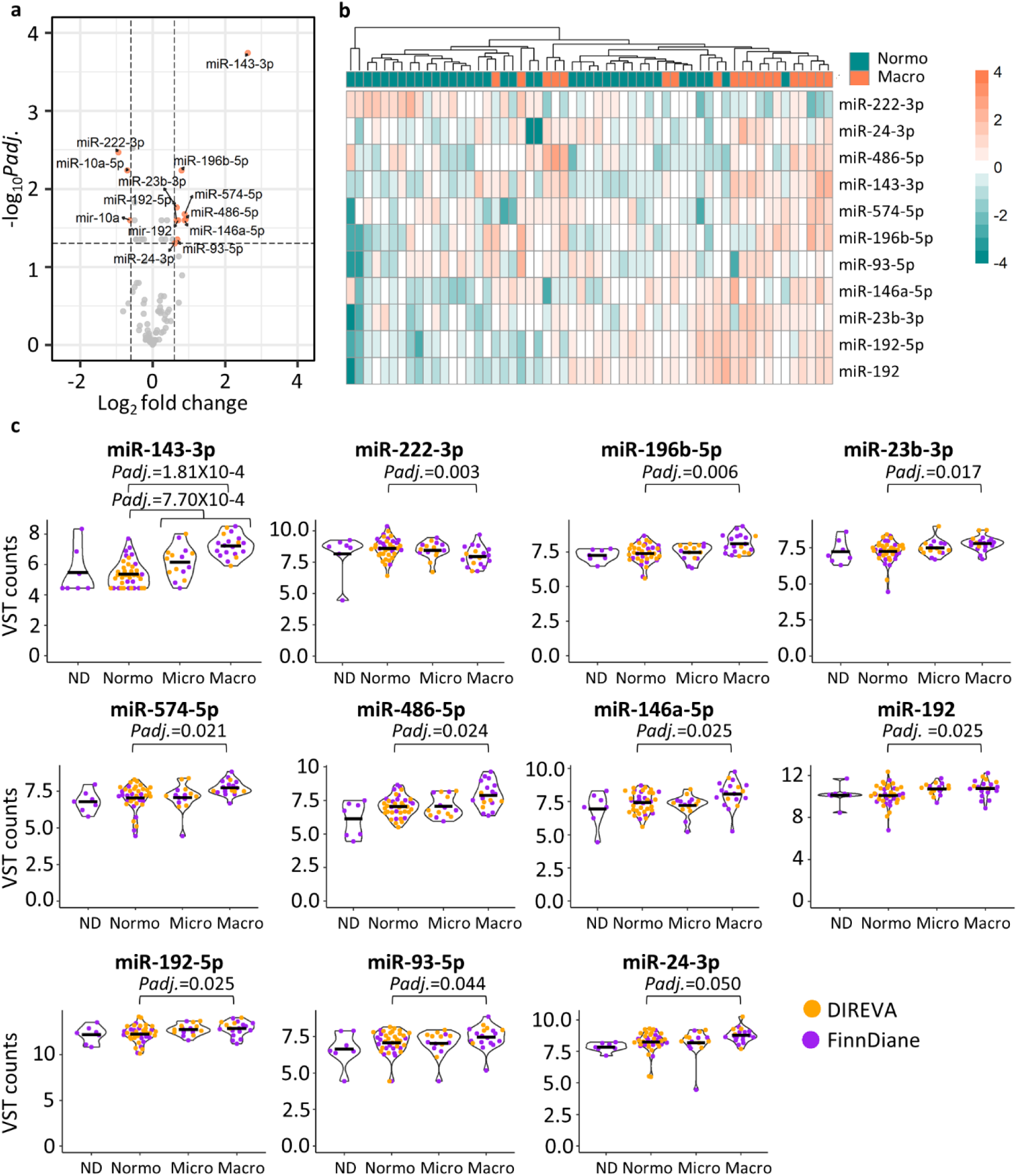
Differential expression analysis of miRNA sequencing data for T1D discovery cohort. Differential expression between normoalbuminuric and macroalbuminuric groups (a-b). (a) Volcano plot presenting differentially expressed miRNAs (mean of five normalized counts, adjusted p-value <0.05, and absolute log2FC >0.6). miR-10a and miR-10a-5p were DE in the preanalytical comparisons. Thus, not considered for further analysis. (b) Heatmap clustering based on the VST normalized read counts of the 11 DE miRNAs. (c) Violin plots from VST normalized data (raw counts were normalized and VST transformed using DeSeq2) including also normo- and microalbuminuria samples. The black horizontal lines show the mean value. Differentially expressed (DE), macroalbuminuria (Macro), microalbuminuria (Micro), non-diabetic control (ND), normoalbuminuria (Normo), adjusted P-value (Padj.), variance stabilizing transformation (VST).

Differential expression analysis between the rest of the groups (Supplementary Table S3c-e) showed significant upregulation of miR-143-3p in normoalbuminuria vs albuminuria comparison (Figure 3c). Non-diabetic individuals were not included in the analyses due to the small sample number and lesser relevance as a control group, but the miRNA expression was similar to the normoalbuminuria group.

### Correlation analysis

We next determined correlations between clinical measurements and the 11 DE miRNAs. Significant correlations were found for miR-143-3p, miR-146a-5p, miR-196b-5p and miR-222-3p (Figure 4a and Supplementary Figure S3).

**Figure 4.**
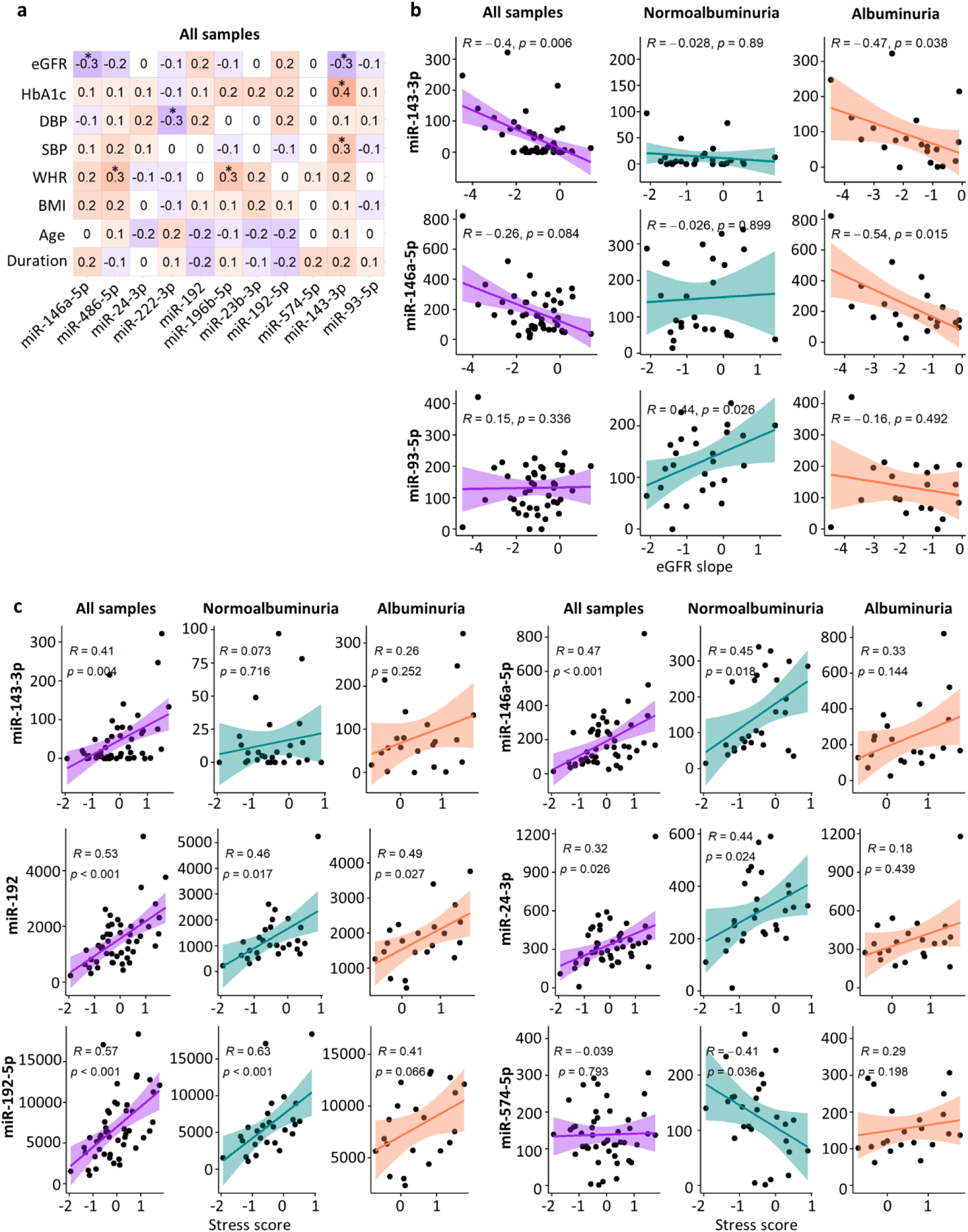
Correlation of normalized miRNA counts with clinical measurements and stress score in the T1D discovery cohort (complete clinical data were available for 69 individuals). Correlations were considered statistically significant if R >0.3 and p <0.05 (a) Spearman correlation of each differentially expressed miRNA and clinical data collected at the time of urine collection. p-value < 0.05 (*) (b) Spearman correlation of eGFR slope data and those miRNAs that included at least one significant p-value. eGFR slope data were available for 46 individuals with T1D (n=26 normo-, 10 micro-, and 10 with macroalbuminuria) (c) Spearman correlation of stress score data and those miRNAs that included at least one significant p-value. Albuminuria group refers to micro- and macroalbuminuria groups combined. The stress score data were available for 48 individuals with T1D (n=27 normo-, 10 micro-, and 11 macroalbuminuria). Body mass index (BMI), diastolic blood pressure (DBP), estimated glomerular filtration rate (eGFR), glycated hemoglobin (HbA1c), macroalbuminuria (Macro), microalbuminuria (Micro), non-diabetic control (ND), systolic blood pressure (SBP), waist-hip ratio (WHR).

To inspect the correlation with the eGFR slope, we analyzed all groups combined, the normoalbuminuria group alone, and the albuminuria group alone. MiR-143-3p correlated significantly with the eGFR slope in all groups combined and in the albuminuria group, miR-146a-5p in the albuminuria group and miR-93-5p in the normoalbuminuria group (Figure 4b).

We additionally analyzed, in the same groups, the correlation with the uEV “stress score”,^15^ elevated in the individuals showing declining kidney function. A significant correlation was found for miR-192 in all three comparisons, and miR-192-5p, miR-146a-5p and miR-24-3p in all groups combined and in the normoalbuminuria group (Figure 4c). MiR-143-3p correlated significantly in all groups combined and miR-574-5p in the normoalbuminuria group (Figure 4c). Together, the DE miRNAs correlated with DKD-associated parameters in different albuminuria groups alone or combined.

### Replication studies

In the T1D replication cohort (Table 1), differential expression was analyzed between the normoalbuminuria vs albuminuria groups. Totally, 11 miRNAs were DE (Figure 5a, Supplementary Table S4). MiR-192-5p, miR-146a-5p, miR-486-5p, and miR-574-5p were similarly DE as in the discovery cohort and not changed in the preanalytical comparisons (Figure 5b). Additionally, miR-192-5p normalized counts correlated significantly with both the eGFR slope and the stress score (Figure 5c, d) and miR-486-5p correlated significantly with the stress score (Figure 5d).

**Figure 5:**
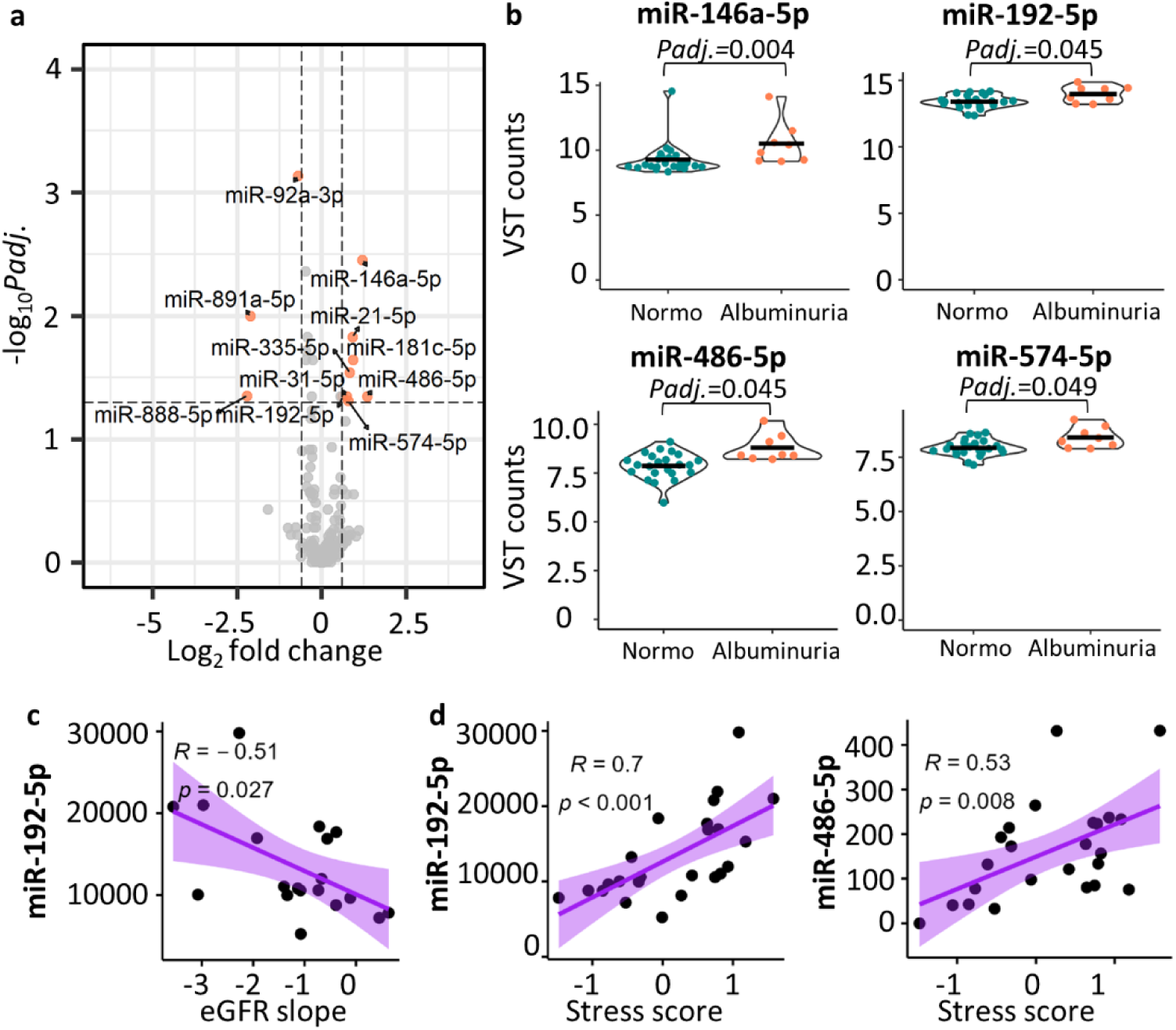
Replication of the miRNA biomarker candidates with an independent T1D female cohort. Sequencing results are shown as (a) Volcano plot of all differentially expressed miRNAs (mean of five normalized counts, adjusted p-value <0.05, and absolute log2FC >0.6) between normoalbuminuria (n=23) vs albuminuria (n=8) groups (micro- and macroalbuminuria) and (b) the expression levels (VST counts, raw counts were normalized and VST transformed using DeSeq2) in violin plots with a black horizontal line marking the mean value for the four differentially expressed miRNAs shared between the T1D discovery and replication cohorts. (c) Spearman correlation between the eGFR slope and miR-192-5p. The eGFR slope was available for 19 individuals (n=16 normo-, 2 micro-, and 1 macroalbuminuria) and (d) Spearman correlation between the stress score and miR-192-5p or miR-486-5p. The stress score data were available for 24 individuals (n=19 normo-, 3 micro-, and 2 macroalbuminuria). Macroalbuminuria (Macro), microalbuminuria (Micro), normoalbuminuria (Normo), P-adjusted value (Padj.), type 1 diabetes (T1D), variance stabilizing transformation (VST).

Differential expression analysis using the T2D replication cohort (Table 1) revealed an upregulation of the miR-192 and miR-192-5p (nominal p-value <0.05) between the macro-vs microalbuminuria groups (Supplementary Table S5).

To further validate the 11 DE miRNAs, we analyzed published uEV miRNA sequencing or qPCR datasets. There, 10 miRNAs presented the same direction of change as in our data (Table 3).

**Table 3.** Published datasets (DKD, CKD or control) used to compare our findings. Macroalbuminuria (Macro), microalbuminuria (micro), non-diabetic (ND), not available (NA), not applicable (N/A), Prostate cancer. * 41% of the individuals with proteinuria and 35% of the individuals without proteinuria had diabetes. ** statistically significant upregulation. From Barreiro et al, 2020 we used the ultracentrifugation dataset.

| Reference | Sample type | miRNA profiling method | Diabetes type or control | Donors (n) | Higher expression in case group | Lower expression in case group | Figure |
| --- | --- | --- | --- | --- | --- | --- | --- |
| Barreiro et al, 2020<sup>23</sup> | uEV | miRNA sequencing (QIAseq miRNA Library, Qiagen) | T1D | ND=5; Macro=5 | miR-146a-5p, miR-192-5p, miR-486-5p, miR-143-3p, miR-196b-5p and miR-23b-3p |  | Supplementary Figure S5a |
| Barutta et al, 2013<sup>22</sup> | uEV | qPCR (Human TaqMan miRNA Array A) | T1D | Normo=2, Micro=2 | miR-143 and miR-23b |  | Supplementary Figure S5b |
| Park et al, 2020 and 2022<sup>25,26</sup> | uEV | miRNA sequencing (SMARTer smRNA-Seq kit, Illumina) | T2D | ND=4 T2D and DKD=5 | miR-146-5p**, miR-192-5p, miR-196b-5p, miR-23-3p | miR-486-5p, miR-93-5p | supplementary Figure S5c |
| Perez-Hernandez et al, 2021<sup>27</sup> | uEV | miRNA sequencing CleanTag Small RNA library preparation kit (TriLink Biotechnologies) | T2D* | Hypertension with albuminuria (UAE) =22; without albuminuria (non-UAE)= 26 | miR-146a-5p, miR-486-5p, miR-93-5p and miR-24-3p | miR-222-3p, miR-192-5p | Supplementary Figure S5d |
| Puhka et al, 2022<sup>24</sup> | uEV | miRNA sequencing (QIAseq miRNA Library, Qiagen) | control | ND= 10; Prostate cancer=30 | N/A | N/A | Supplementary Figure S5e |

To assess whether the DE miRNAs show specificity for DKD we studied the prostate cancer sequencing dataset from Puhka et al, 2022. Hierarchical clustering analysis of 10 miRNAs (detected) showed lack of clusters by any disease or age status (Supplementary Figure S5e). The replication studies thus supported changes in the candidate biomarker miRNAs in DKD.

### Validation by qPCR

UEV miss stable reference miRNAs for qPCR-based validation. We previously generated a candidate list of 14 stable uEV miRNAs ^31^ that we now evaluated further; we checked whether they were DE in the preanalytical and DKD analyses here. 6 candidate miRNAs remained stable in all the comparisons, supporting their use as reference miRNAs (Table 4).

**Table 4:**
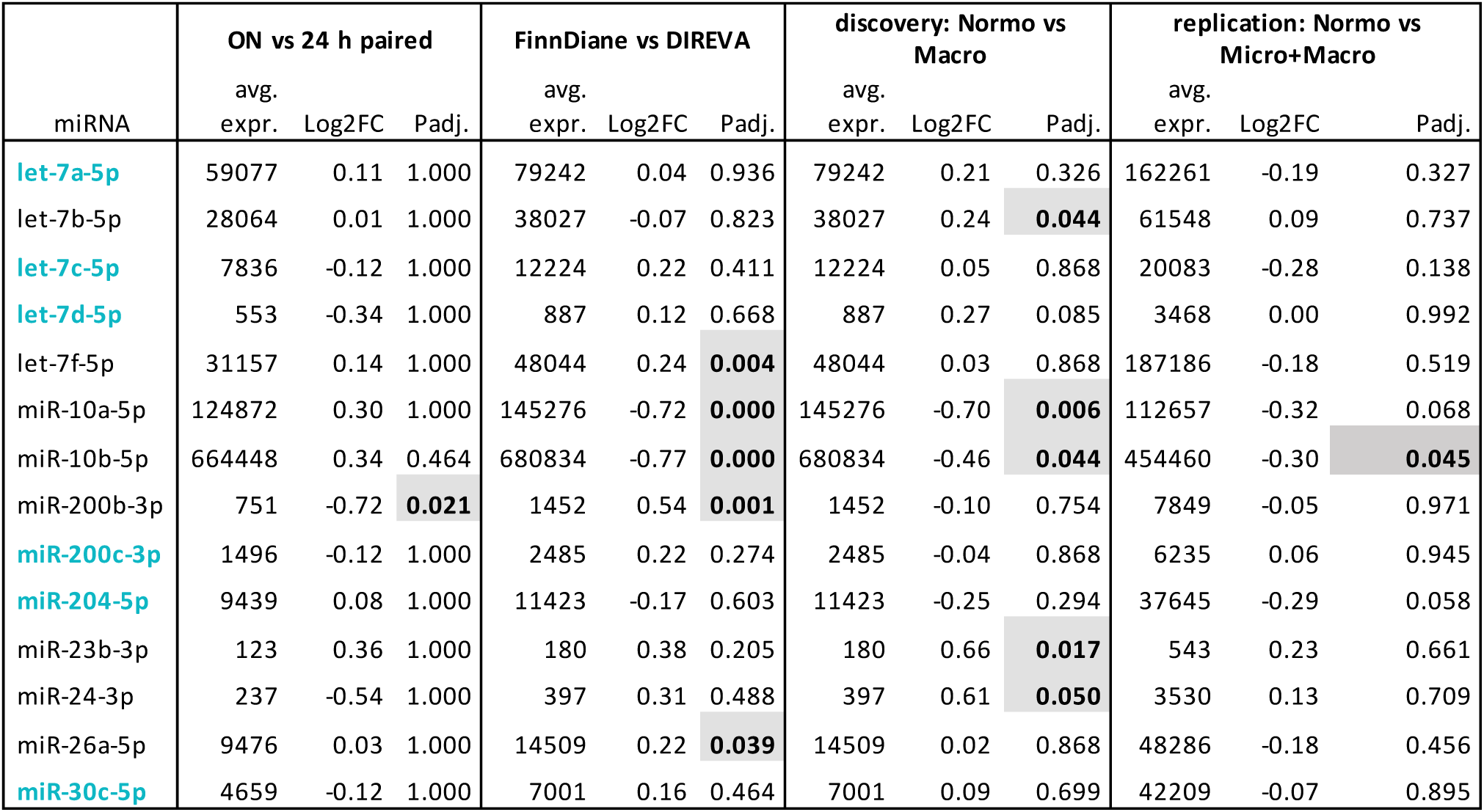
Evaluation of 14 reference miRNA candidates across the preanalytical urine collection and T1D DKD datasets. Six miRNAs (blue) remained stable and eight (grey highlight) showed differential expression in some of the comparisons. Fold change (FC), macroalbuminuria (Macro), microalbuminuria (Micro), normoalbuminuria (Normo), overnight (ON), p-adjusted value (padj.).

| miRNA | ON vs 24 h paired |  |  | FinnDiane vs DIREVA |  |  | discovery: Normo vs Macro |  |  | replication: Normo vs Micro+Macro |  |  |
| --- | --- | --- | --- | --- | --- | --- | --- | --- | --- | --- | --- | --- |
|  | avg. expr. | Log2FC | Padj. | avg. expr. | Log2FC | Padj. | avg. expr. | Log2FC | Padj. | avg. expr. | Log2FC | Padj. |
| let-7a-5p | 59077 | 0.11 | 1.000 | 79242 | 0.04 | 0.936 | 79242 | 0.21 | 0.326 | 162261 | -0.19 | 0.327 |
| let-7b-5p | 28064 | 0.01 | 1.000 | 38027 | -0.07 | 0.823 | 38027 | 0.24 | **0.044** | 61548 | 0.09 | 0.737 |
| let-7c-5p | 7836 | -0.12 | 1.000 | 12224 | 0.22 | 0.411 | 12224 | 0.05 | 0.868 | 20083 | -0.28 | 0.138 |
| let-7d-5p | 553 | -0.34 | 1.000 | 887 | 0.12 | 0.668 | 887 | 0.27 | 0.085 | 3468 | 0.00 | 0.992 |
| let-7f-5p | 31157 | 0.14 | 1.000 | 48044 | 0.24 | **0.004** | 48044 | 0.03 | 0.868 | 187186 | -0.18 | 0.519 |
| miR-10a-5p | 124872 | 0.30 | 1.000 | 145276 | -0.72 | **0.000** | 145276 | -0.70 | **0.006** | 112657 | -0.32 | 0.068 |
| miR-10b-5p | 664448 | 0.34 | 0.464 | 680834 | -0.77 | **0.000** | 680834 | -0.46 | **0.044** | 454460 | -0.30 | **0.045** |
| miR-200b-3p | 751 | -0.72 | **0.021** | 1452 | 0.54 | **0.001** | 1452 | -0.10 | 0.754 | 7849 | -0.05 | 0.971 |
| miR-200c-3p | 1496 | -0.12 | 1.000 | 2485 | 0.22 | 0.274 | 2485 | -0.04 | 0.868 | 6235 | 0.06 | 0.945 |
| miR-204-5p | 9439 | 0.08 | 1.000 | 11423 | -0.17 | 0.603 | 11423 | -0.25 | 0.294 | 37645 | -0.29 | 0.058 |
| miR-23b-3p | 123 | 0.36 | 1.000 | 180 | 0.38 | 0.205 | 180 | 0.66 | **0.017** | 543 | 0.23 | 0.661 |
| miR-24-3p | 237 | -0.54 | 1.000 | 397 | 0.31 | 0.488 | 397 | 0.61 | **0.050** | 3530 | 0.13 | 0.709 |
| miR-26a-5p | 9476 | 0.03 | 1.000 | 14509 | 0.22 | **0.039** | 14509 | 0.02 | 0.868 | 48286 | -0.18 | 0.456 |
| miR-30c-5p | 4659 | -0.12 | 1.000 | 7001 | 0.16 | 0.464 | 7001 | 0.09 | 0.699 | 42209 | -0.07 | 0.895 |

We confirmed 4 candidate reference miRNAs by qPCR. Let-7a-5p and miR-200c-3p gave the best correlation i.e., similar stability and were used for normalization of 6 DE miRNAs (Supplementary results and Figure 6). In the T1D discovery cohort, qPCR confirmed a significant difference for the normo-vs macroalbuminuria and normo-vs albuminuria groups for miR-143-3p, miR-192-5p, miR-222-3p, miR-23b-3p, and miR-24-3p (Figure 6g-k, Supplementary Table S6a, b). For the T1D replication cohort, miR-192-5p showed significant differences between the normoalbuminuria and albuminuria groups (Figure 6m, Supplementary Table S6c, d)

**Figure 6:**
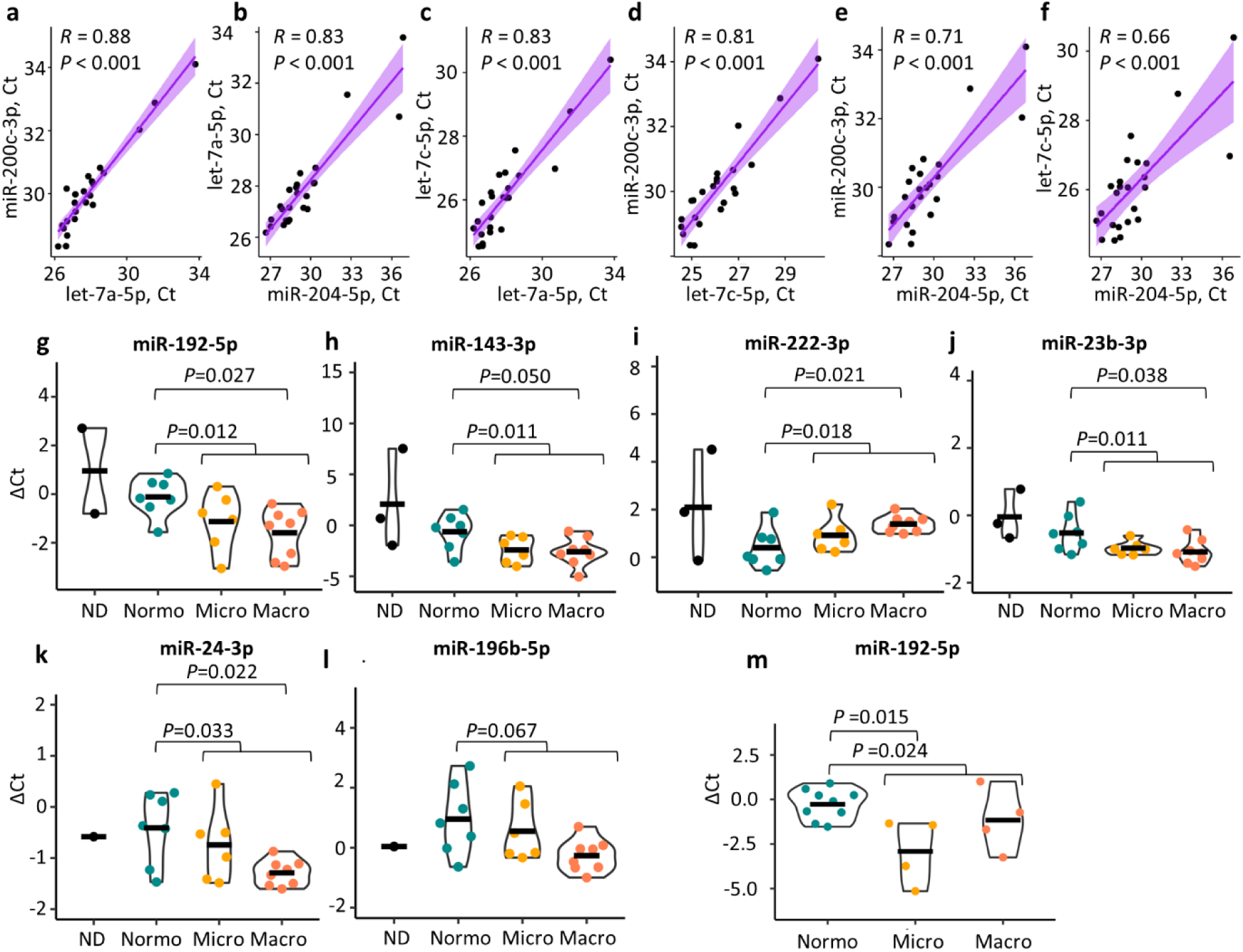
Quantitative PCR validation of selected miRNAs in the T1D discovery cohort. A subset of 21 individuals with T1D (n= 7 normo-, 6 micro-, and 8 macroalbuminuria) and 3 non-diabetic individuals were included. (a,f) Spearman correlation of the cycle threshold (Ct) values between the candidate normalization miRNAs. (g-l) Delta Ct data for the candidate biomarker miRNAs represented in violin plots. The black horizontal line shows the mean value. (m) Quantitative PCR of miR-192-5p for 17 individuals from the T1D replication cohort (n=9 normo-vs 4 micro- and 4 macroalbuminuria combined). Delta Ct values were calculated relative to the geometric mean of let-7a-5p and miR-200c-3p. MiRNAs were considered significantly DE with p values <0.05. Macroalbuminuria (Macro), microalbuminuria (Micro), non-diabetic control (ND) normoalbuminuria (Normo).

### ROC curve analysis

Kidney dysfunction can be measured by eGFR decline, but measurement of a slope requires a long time for robust estimates. To assess whether the candidate miRNAs could be used to identify the fastest eGFR decliners (lowest quartile, Q1), we performed ROC analysis. We first studied individual miRNAs and some typical clinical measurements (eGFR – expressed as 1/eGFR, HbA1c, SBP). We plotted the 4 miRNAs validated in the T1D female cohort (miR-146a-5p, miR-192-5p, miR-486-5p, miR-574-5p) for all cohorts, and miR-143-3p (highest FC and adjusted p-value in the discovery cohort), for the discovery cohort. The best performing miRNAs (area under the curve, AUC > 0.7) were miR-143-3p and miR-146a-5p for the T1D discovery cohort, miR-192-5p and miR-146a-5p for the T1D replication cohort and miR-486-5p for the T2D replication cohort (Figure 7a-c, Supplementary results). Clinical measurements presented generally lower AUC values than best performing miRNAs (Figure 7d-f), except eGFR that produced an AUC of 1 for the T1D replication cohort (Figure 7e).

**Figure 7:**
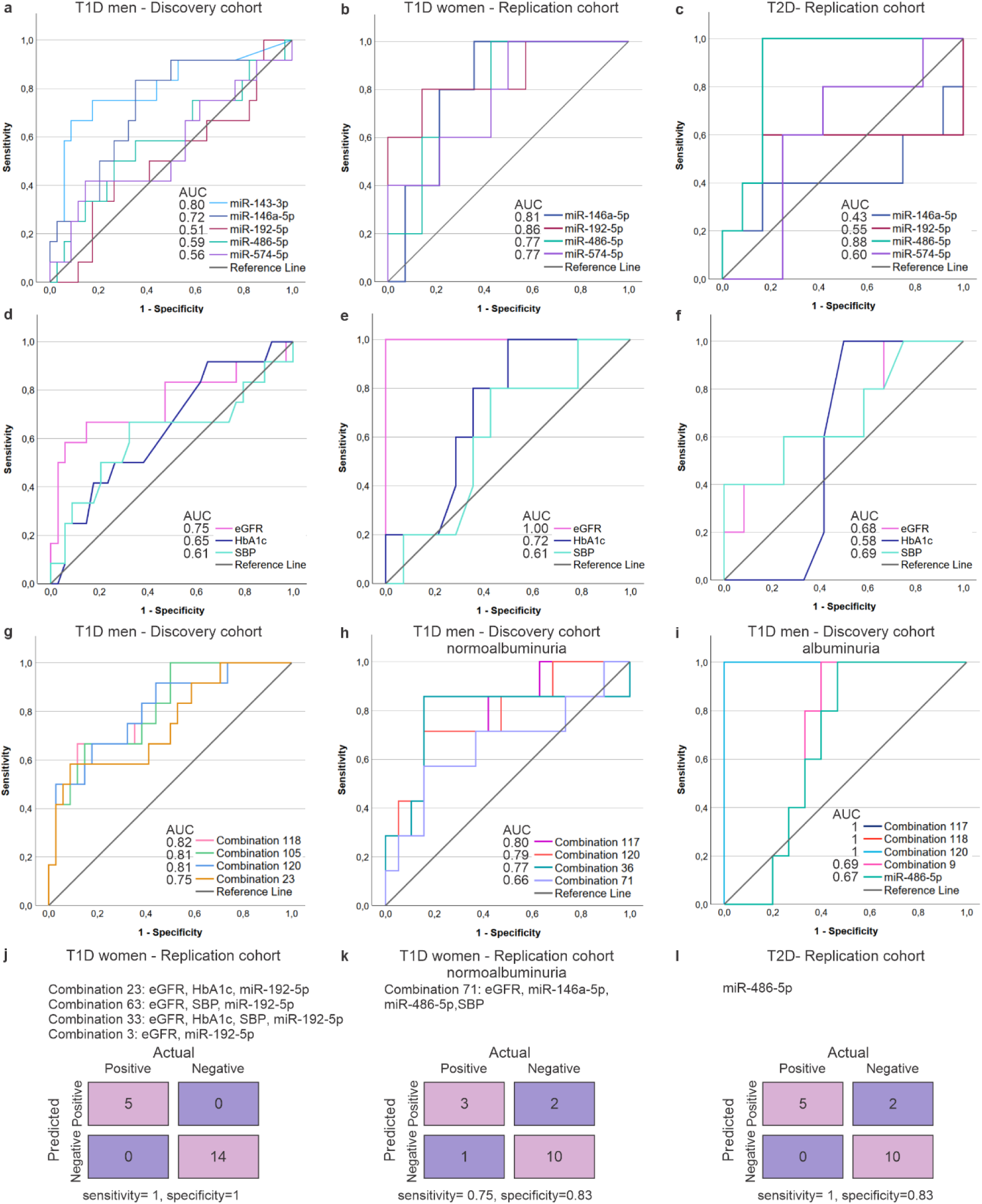
ROC (receiver operator characteristic) analysis and confusion charts using sequencing and clinical data. The number of individuals with available eGFR decline data has been indicated for each cohort (a-f) ROC curves of selected miRNAs and clinical parameters in discovery and replication cohorts for predicting eGFR decline (lowest quartile, Q1) of (a,d) ≤-1,59 mL/min/1.73 m2/year for T1D male discovery cohort (n= 46, 16 normo-, 10 micro-, and 10 macroalbuminuria), (b,e) ≤-1,66 mL/min/1.73 m2/year for T1D female replication cohort (n= 19, 16 normo-, 2 micro-, and 1 macroalbuminuria) and (c-f) ≤-3.08 mL/min/1.73 m2/year for T2D replication cohort (n= 17, 10 micro-, and 7 macroalbuminuria). (g-i) ROC curves of selected combinations of validated miRNAs in T1D cohorts (miR-146a-5p, miR-192-5p, miR-486-5p, miR-574-5p) and clinical measurements computed using CombiROC R package for predicting eGFR decline (lowest quartile Q1) of (g) same as in a,d, (h) ≤-1,36 mL/min/1.73 m2/year for normoalbuminuric subgroup of T1D discovery cohort and (i) ≤-2,46 mL/min/1.73 m2/year for albuminuric subgroup of T1D discovery cohort. (j-l) Results of the best classifications of replication cohorts using the models generated with CombiROC for predicting eGFR decline (lowest quartile, Q1) of (j) same as b,e, (k) ≤-1,18 mL/min/1.73 m2/year for normoalbuminuric subgroup of T1D replication cohort, and (l) same as c-f. The compositions of the combinations are disclosed in Supplementary Table S7. Area under the curve (AUC), Estimated glomerular filtration rate (eGFR, used as 1/eGFR), glycated hemoglobin (HbA1c), systolic blood pressure (SBP), type 1 diabetes (T1D), type 2 diabetes (T2D)

The miRNAs and clinical variables were next combined to evaluate whether ROC results could be improved and to generate models to test the classification of the replication cohorts. For this combinatorial analysis, we used the T1D discovery cohort (our largest cohort) for 3 analyses to classify the fastest eGFR decliners including i) all the samples, ii) the normoalbuminuria subgroup, and iii) the albuminuria (micro- and macroalbuminuria) subgroup (Figures 7f-h, Supplementary Tables S7a-c). For all 3 analysis we found combinations with AUC ≥0.80.

To identify the best models to classify the fastest eGFR decliners in the replication cohorts, we selected from each combinatorial and individual miRNA analysis (i, ii, iii) the models with an AUC>0.65 and applied those models to the replication cohorts (Supplementary Tables S8a-c). The models with the best specificity and sensitivity are shown in Figure 7j-l. Altogether, with 0-2 false positives or negatives, the models showed the potential of uEV miRNAs for improved evaluation of kidney function from a single urine sample.

### Target pathway, mRNA and protein analysis

To understand which pathways the DE miRNAs modulate in DKD, we studied mRNA targets expressed in the kidney (Supplementary Table S9) – 9 of the miRNAs had 54 experimentally validated mRNA targets. To simplify their target pathways, we grouped the various diseases and functions into larger categories (Supplementary Table S10). From the 26 categories, 16 were directly DKD-associated. (Figure 8). The top DKD-associated categories (as judged by the number of incoming links/regulations) included kidney hypertrophy; kidney injury; apoptosis; and immune and inflammatory response. Other strongly DKD-associated categories - such as glomerulosclerosis, albuminuria, fibrosis, and reactive oxygen species (ROS) production - were represented but with less mRNA targets. To obtain more evidence for the relevance of the targeted mRNAs and pathways in DKD, we incorporated kidney single cell sequencing data from individuals with T2D and DKD^32^. We identified 19 cell-type categorized mRNA targets that were DE in DKD biopsies. The cells with most of the dysregulated mRNAs (number of incoming links) were endothelial cells, thick ascending limb of Henle’s loop CLDN16 (-), podocytes, and fibroblast.

**Figure 8:**
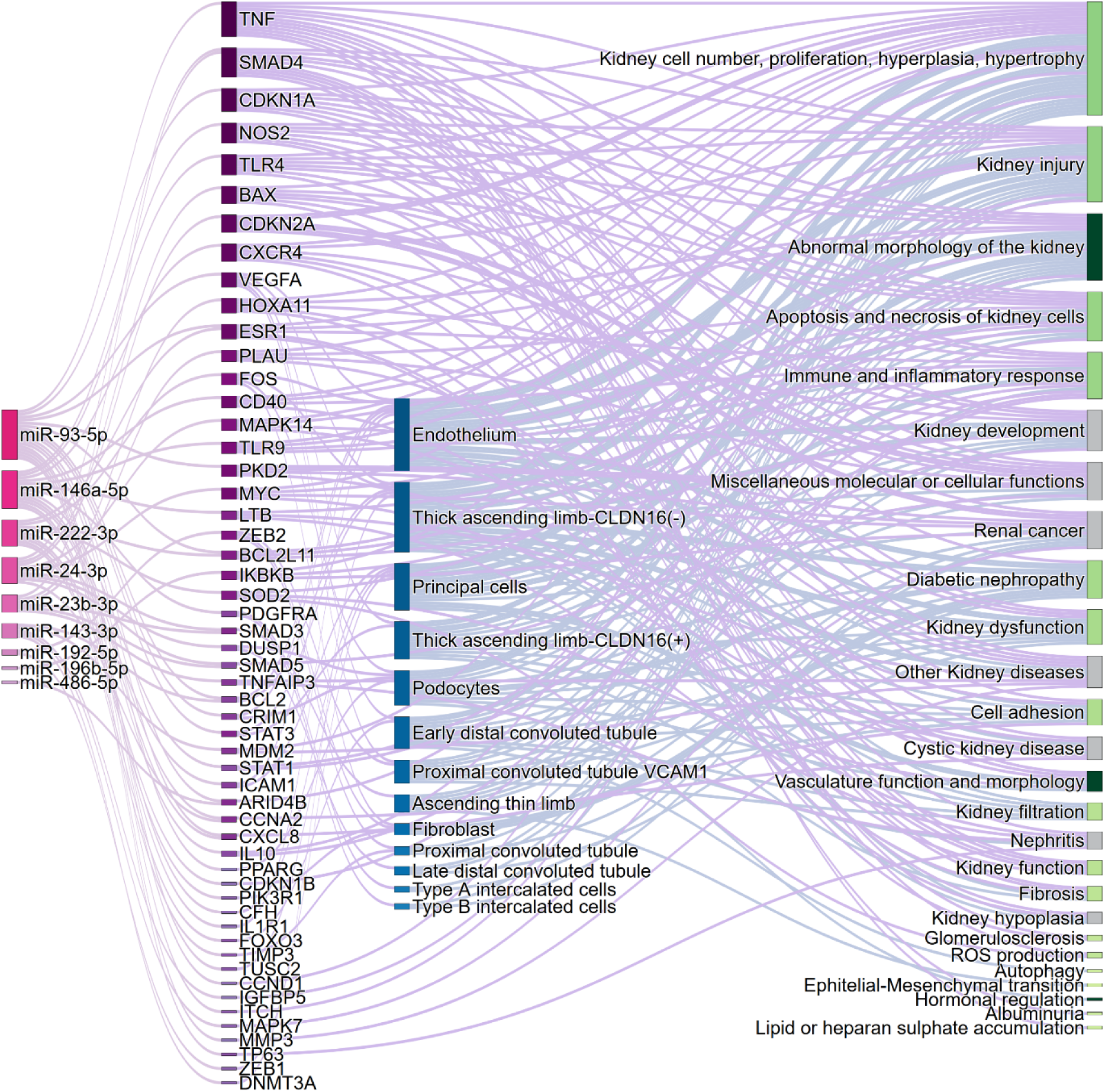
Interactive sankey diagram depicting differentially expressed miRNAs between macro- and normoalbuminuria groups in the T1D discovery cohort, their experimentally validated target mRNAs, cell types where the targets have been found to be differentially expressed in kidney biopsies from T2D and DKD (scRNA seq data, Wilson et al, 2022) and disease or function associated with the target mRNAs. Light green nodes are associated with DKD, dark green nodes are associated with DKD but also with physiological conditions, and grey nodes are not associated with DKD. Source and target nodes are also disclosed in Supplementary Table S11. Diabetic kidney disease (DKD), reactive oxygen species (ROS), RNA single cell sequencing (scRNA), type 1 diabetes (T1D), type 2 diabetes (T2D). Additionally, a predicted target search showed many other interesting targets linked to DKD e.g., transcription factors HNF1A, HNF1B, HNF4A, SMAD2, and STAT6 (Supplementary table S9).

Additionally, a predicted target search showed many other interesting targets linked to DKD e.g., transcription factors HNF1A, HNF1B, HNF4A, SMAD2, and STAT6 (Supplementary table S9).

We further studied whether the proteins coded by the 54 target mRNAs (see Figure 8) or by the DE uEV mRNAs (including the stress score) we identified in DKD^15^, were dysregulated in individuals with T1D and T2D. We had a large plasma proteomics dataset available in the UK biobank that were compared in individuals with and without kidney or ophthalmic diabetes complications. Males and females were analyzed both together and separately when possible. Data were available for 34 proteins. Logistic regression model analysis showed a statistically significant association with kidney complications for 9 proteins (3 in common with ophthalmic complications) in the combined and male cohorts (Table 5).

**Table 5:** Statistically significant associations from the UK biobank plasma proteomic analysis using logistic regression models. *MiRNA’s predicted with high or moderate confidence (Supplementary Table S9) – miRNA’s without star have target with experimental evidence, kidney complications (KC), ophthalmic complications (OC), p-adjusted value (padj.), standard error (SEM), type 1 diabetes (T1D), type 2 diabetes (T2D).

| protein | miRNA | strata | case | contro<br>l | n<br>cases | n<br>controls | $\beta$ | SE<br>M | z-<br>score | Padj. |
| --- | --- | --- | --- | --- | --- | --- | --- | --- | --- | --- |
| BCL2 | miR-143-3p, miR-93-5p,<br>miR-23b-3p* | combined | T2D<br>KC | T2D | 85 | 3822 | 0.3<br>1 | 0.0<br>6 | 4.83 | 1.41E-04 |
| BCL2 | miR-143-3p, miR-93-5p,<br>miR-23b-3p* | males | T2D<br>KC | T2D | 60 | 2271 | 0.3<br>1 | 0.0<br>8 | 3.92 | 9.22E-03 |
| BCL2L1<br>1 | miR-222-3p, miR-93-5p,<br>miR-24-3p* | males | T2D<br>KC | T2D | 60 | 2271 | 0.6<br>4 | 0.1<br>7 | 3.83 | 1.29E-02 |
| BCL2L1<br>1 | miR-222-3p, miR-93-5p,<br>miR-24-3p* | combined | T2D<br>KC | T2D | 85 | 3822 | 0.4<br>8 | 0.1<br>4 | 3.52 | 4.44E-02 |
| BCL2L1<br>1 | miR-222-3p, miR-93-5p,<br>miR-24-3p* | combined | T2D<br>OC | T2D | 435 | 3510 | 0.2<br>2 | 0.0<br>6 | 3.84 | 1.26E-02 |
| CRIM1 | miR-93-5p | combined | T2D<br>KC | T2D | 85 | 3822 | 0.5<br>4 | 0.0<br>8 | 7.07 | 1.64E-10 |
| CRIM1 | miR-93-5p | males | T2D<br>KC | T2D | 60 | 2271 | 0.6<br>7 | 0.1<br>0 | 7.01 | 2.44E-10 |
| IL1R1 | miR-146a-5p | combined | T2D<br>KC | T2D | 85 | 3822 | 0.5<br>5 | 0.0<br>8 | 7.14 | 9.51E-11 |
| IL1R1 | miR-146a-5p | males | T2D<br>KC | T2D | 60 | 2271 | 0.6<br>2 | 0.0<br>9 | 6.89 | 5.71E-10 |
| IL1R1 | miR-146a-5p | combined | T2D<br>OC | T2D | 435 | 3510 | 0.3<br>5 | 0.0<br>4 | 8.75 | 2.22E-16 |
| IL1R1 | miR-146a-5p | males | T2D<br>OC | T2D | 283 | 2067 | 0.3<br>4 | 0.0<br>5 | 6.94 | 4.03E-10 |
| IL1R1 | miR-146a-5p | females | T2D<br>OC | T2D | 152 | 1443 | 0.3<br>6 | 0.0<br>7 | 5.36 | 8.61E-06 |
| MMP3 | miR-93-5p, miR-574-5p* | combined | T2D<br>KC | T2D | 85 | 3822 | 0.5<br>5 | 0.1<br>0 | 5.48 | 4.30E-06 |
| MMP3 | miR-93-5p, miR-574-5p* | males | T2D<br>KC | T2D | 60 | 2271 | 0.6<br>6 | 0.1<br>2 | 5.34 | 9.69E-06 |
| PDGFR<br>A | miR-146a-5p, miR-24-3p*,<br>miR-93-5p* | combined | T1D<br>KC | T1D | 12 | 416 | 0.8<br>3 | 0.2<br>3 | 3.65 | 8.89E-03 |
| PDGFR<br>A | miR-146a-5p; miR-24-3p*,<br>miR-93-5p* | combined | T2D<br>KC | T2D | 85 | 3822 | 0.5<br>9 | 0.0<br>8 | 7.19 | 6.77E-11 |
| PDGFR<br>A | miR-146a-5p, miR-24-3p*,<br>miR-93-5p* | males | T2D<br>KC | T2D | 60 | 2271 | 0.6<br>5 | 0.1<br>0 | 6.65 | 3.10E-09 |
| PDGFR<br>A | miR-146a-5p, miR-24-3p*,<br>miR-93-5p* | combined | T1D<br>OC | T1D | 90 | 354 | 0.4<br>6 | 0.1<br>1 | 4.41 | 1.07E-03 |
| PDGFR<br>A | miR-146a-5p, miR-24-3p*,<br>miR-93-5p* | males | T1D<br>OC | T1D | 48 | 200 | 0.6<br>6 | 0.1<br>5 | 4.39 | 1.15E-03 |
| PDGFR<br>A | miR-146a-5p, miR-24-3p*,<br>miR-93-5p* | combined | T2D<br>OC | T2D | 435 | 3510 | 0.1<br>7 | 0.0<br>5 | 3.80 | 1.51E-02 |
| RBP5 | miR-192-5p*, miR-574-5p* | males | T2D<br>KC | T2D | 60 | 2271 | 0.5<br>1 | 0.1<br>1 | 4.54 | 5.86E-04 |
| RBP5 | miR-192-5p*, miR-574-5p* | combined | T2D<br>KC | T2D | 85 | 3822 | 0.4<br>0 | 0.0<br>9 | 4.32 | 1.58E-03 |
| TNF | miR-93-5p, miR-24-3p* | combined | T2D<br>KC | T2D | 85 | 3822 | 0.3<br>8 | 0.0<br>6 | 6.65 | 2.96E-09 |
| TNF | miR-93-5p, miR-24-3p* | males | T2D<br>KC | T2D | 60 | 2271 | 0.4<br>3 | 0.0<br>7 | 5.86 | 4.69E-07 |
| VEGFA | miR-93-5p | males | T2D<br>KC | T2D | 60 | 2271 | 0.5<br>7 | 0.1<br>2 | 4.80 | 1.65E-04 |
| VEGFA | miR-93-5p | combined | T2D<br>KC | T2D | 85 | 3822 | 0.4<br>6 | 0.1<br>0 | 4.57 | 4.90E-04 |

In summary, all target analyses supported the relevance of the miRNA candidates in DKD.

## Discussion

The uEV provide a view of the kidney tissue, which is rarely biopsied in individuals with diabetes. Relatively few studies have profiled uEV miRNAs in DKD. Common shortcomings regarding the uEV miRNA studies are poor characterization of the uEV isolates, low statistical power, heterogenous groups, and lack of replication in independent cohorts. Here, we used several well characterized T1D, T2D and control cohorts to evaluate potential uEV-miRNA changes in DKD. We found candidate DKD marker miRNAs, which regulate diverse DKD-associated RNAs, proteins and pathways and could classify the individuals with the fastest eGFR decline in early to late DKD.

MiRNA biomarker candidates tend to differ between studies, not least due to preanalytics.^13,18,20^ We therefore started by exploring our preanalytical variables, which revealed some DE miRNAs between the overnight vs 24 h urine collections. We then selected only the unaffected miRNAs for further analysis of the DE and reference miRNAs – the safest strategy when including different cohorts in the same study.

Pathway analysis of the DE miRNAs revealed their overall relevance in DKD: many target mRNAs have been linked to DKD^33–37^ and they are part of the classical DKD pathways.^3,10,38^ Interestingly, judged by the number of targets per disease pathway, uEV captured proportionally more changes at the early DKD stage, such as kidney hypertrophy, kidney injury, and changes in the kidney morphology e.g., expansion of mesangial matrix^39,40^. Other hallmark pathways, such as autophagy and ROS production,^38^ and advanced DKD changes like glomerulosclerosis and fibrosis,^39,40^ were less targeted by the DE miRNAs. Incorporation of single cell sequencing data from human kidney biopsies with DKD^32^ showed that many mRNA targets were DE in the hallmark kidney cell types of DKD, such as the endothelium, podocytes, and proximal tubule cells.^39^ Since the biopsies were taken at the early stages of DKD,^32^ it is possible that our DE miRNAs (that regulate the DE biopsy mRNAs) also relate to the early changes in DKD. Additionally, exploration of the proteins encoded by the DE miRNAs’ target mRNAs and the stress score mRNAs revealed nine proteins associated with kidney and ophthalmic complications in independent T1D and T2D studies. Finding such a large proportion of target proteins (26%) changed in plasma and prior data of protein changes in DKD or CKD - e.g., for plasma VEGF,^41^ plasma IL1R1,^42^ plasma and podocyte CRIM1,^43^ tubular and podocyte BCL2,^44,45^ plasma and serum MMP3,^46,47^ and plasma TNF^48^ - support the hypothesis that the DE miRNAs are important regulators in DKD.

The 4 validated miRNAs in DKD of T1D showed individually and in combinations good performance in the ROC analyses and classifications for the T1D or T2D replication cohorts. From these candidate DKD markers, the miR-192-5p was consistently upregulated in the discovery and many replication cohorts and correlated with the eGFR slope and the stress score. In line, prior studies have shown miR-192-5p upregulation in urine^18,49–51^ and correlation between its expression in uEV and urinary sediments and the eGFR in individuals with T2D and DKD.^19,49^ Early on, the miR-192 was studied in human kidney cortical tissues and in glomeruli of rat and mouse models of DKD.^50^ The miR-192 upregulation in the kidneys could link to e.g., the inhibition of GLP-1 receptor or SMAD/TGF-β and PTEN/PI3K/AKT signaling pathways that regulate epithelial-to-mesenchymal transition and fibrosis.^50^ Interestingly, Chatterjee and colleagues^52^ reported that miR-192-5p, miR-146-5p and miR-21-5p - targeting the SMAD/TGF-β pathway - were changed in blood EVs from individuals with the cardiovascular-kidney-metabolic syndrome. Treating proximal tubule cells on a chip with these plasma EVs induced kidney injury markers. As the exact same miRNAs were changed in our DKD study (even if miR-21-5p was excluded due to changes in the preanalytical study), it suggests that for diseases with cardiorenal complications, kidney injury coincides with the secretion of blood and uEV carrying these miRNAs.

The association of miR-146a-5p with diabetes complications have been proven by many studies.^53^ However, the reports have been contradictory e.g., with tissue up- or downregulation and pro- or anti-inflammatory roles in human T2D kidney biopsies and rodent diabetes models^54–56^. Furthermore, uEV miR-146a-5p was found to be upregulated in both lupus nephritis,^57^ and in DKD (here). Studies on uEV cargo sorting are ongoing and could apply active or passive mechanisms serving for (disease-specific) cargo functions or disposal.^58,59^ MiRNAs are also versatile (post-) transcriptional regulators^60 61,62^, andknown as repressors, or rarely, activators of translation.^63^ Consequently, particular miRNAs could be dysregulated in diverse diseases, e.g., we previously observed upregulation of the miR-146a-5p in uEV from advanced prostate cancer groups.^24^ Nevertheless, a combination of 10 DE miRNAs from our DKD discovery study - including the miR-146a-5p - did not separate any prostate cancer study groups. Further, our analysis of the published DKD datasets supported changes in 10 DE miRNAs including miR-146a-5p, miR-192-5p and miR-486-5p. When searching for replication datasets, we encountered difficulties related to data availability and reporting. We failed to find any uEVs studies that had compared individuals with diabetes and normo-vs macroalbuminuria, as we had done to discover our main findings. Thus, it is plausible that the different study designs of the published datasets could explain why some miRNAs showed smaller changes than in our data. Taken together, a larger combination of miRNAs (and clinical variables) has the potential to provide DKD-specificity.

Mir-574-5p has been linked to immune and inflammatory responses, e.g., its overexpression in DKD downregulated C7 in mesangial cells^64^. The DKD-association of miR-486-5p is suggested by its downregulation in glomeruli and proximal tubule cells of biopsies from individuals with DKD^65^ and in mesangial cells exposed to high glucose, which promotes fibrosis.^66^ Interestingly, miR-486-5p is the most abundant miRNA in plasma ^67^ and this opens up the question whether the increased uEV miR-486-5p in DKD could originate from the blood. In healthy conditions, the kidney filtration barrier limits the passage from blood to urine, but when the barrier is disrupted, like in DKD, the passage could increase. ^13,68^ However, to our understanding, more evidence is needed to clarify this phenomenon and its magnitude.

We foresee different potential use of the validated miR-146a-5p, miR-192-5p, miR-486-5p and miR-574. Because they all performed fine as part of combinations to discriminate the fastest eGFR decliners in the T1D normoalbuminuria groups, they might possess the power to serve as early DKD biomarkers. The latter three performed fine also for the albuminuria groups suggesting further use as markers at late stages. In contrast, in the T2D cohort, only the miR-486-5p performed satisfactorily. The difference between the T1D and T2D cohorts was expected based on the different disease etiologies, T2D heterogeneity and co-morbidities.^9,39,69^ However, having found one miRNA (miR-486-5p) in common between the cohorts shows that some prominent mechanisms are shared between the sexes and the diabetes types 1 and 2.

Along with the common DE miRNAs of the T1D male and female cohorts, we discovered some unique miRNAs. Interestingly, two miRNAs DE only in males, target HNF4A (miR-143-3p) or KDM6A (miR-23b-3p), which were recently linked to a sex-specific course of DKD^8^. HNF4A, known from maturity-onset diabetes of the young (MODY) Type 1, regulates kidney metabolism^70^ with androgen receptor particularly in the proximal tubule cells^8,71,72^. KDM6A is a X-linked gene and an epigenetic regulator of genes and metabolism in the kidneys and proximal tubule cells. Also DE in males only, miR-24-3p^73^ targets further MODY genes, HNF1A^74^ and HNF1B^75^, which associate with the regulation of sodium/glucose cotransporter 2 in proximal tubule cells^76^ and kidney disease susceptibility^75^, respectively. Like us, Argyropoulos and colleagues^77^ observed miR-23b-3p and miR-192-5p changes between sexes and in normoalbuminuric individuals subsequently developing microalbuminuria. Of note, while miR-192-5p was DE in both sexes in our study, it classified the fastest eGFR decliners better in females. MiR-222-3p, another DE miRNA only in male uEV, is X-linked,^78^ which could partly explain its sex-specific regulation in DKD. Finally, the plasma proteins targeted by the DE miRNAs (discovered in males) associated with DKD only in male and combined sex cohorts in the UK Biobank. In line, upregulation of plasma VEGF (target of miR-93-5p, DE only in males) was found earlier only in males with DKD^41^. Plasma/serum MMP3 (target of miR-93-5p, and miR-574-5p, DE in both sexes) was associated with eGFR decline^47^ or macroalbuminuria and diabetic retinopathy^46^ in both sexes. While the small size of our female T2D kidney complication group certainly affected the statistical power, the female T2D ophthalmic complication group was large. Since diabetic retinopathy is a strong predictor of DKD,^79^ some shared protein changes can be expected in both diagnoses. However, only IL1R1 (target of miR-146a-5p, DE in both sexes) showed changes in the female T2D ophthalmic complication group along with all male and combined groups. Put against this background, the DE miRNAs appear to target key metabolic and other kidney regulators thereby boosting DKD progression in sex-specific and sex-independent ways. For maximally specific or generally applicable DKD biomarkers, our results support the idea to study uEV both separately and together in males and females.

The strengths of our work include focus on T1D, high-quality samples and characterization of the participants, a long follow-up time, and replication of the results in several independent cohorts. We present a balanced experimental design regarding albuminuria groups and clinical characteristics especially for the discovery cohort. We generated a unique insight into the technical/biological replicability and to disease, diabetes subtype, sex, mRNA, protein, pathway, and kidney cell type specificity. Moreover, we characterized our uEV preparations (here and previously;^15,21,23,31,80^) with many current techniques following MISEV guidelines^81^. The sample number is high compared with previous uEV small-RNA sequencing studies. However, we recognize that the study size is still small for a biomarker study. Thus, our results should be validated in larger and yet more varied cohorts.

In conclusion, we discovered non-invasive candidate miRNA markers for DKD in individuals with T1D. The miRNAs hold the potential to serve as early biomarkers for DKD, because they can modulate early DKD pathways and classify T1D individuals with normoalbuminuria and fast eGFR decline. Our results support that combinations of miRNAs and clinical variables could increase the diagnostic power. Additionally, we advance the uEV liquid biopsy field by pinpointing candidate reference miRNAs and miRNAs affected by common pre-analytical variables.

## Data Availability

The raw count data for miRNA sequencing data used in this manuscript (discovery phase - T1D FinnDiane and DIREVA studies) is provided in the Supplementary Table S3a.
Any additional information required to reanalyze the data reported in this paper is available from the lead contact upon request compatible with the study consents.

## Data sharing statement

The raw count data for miRNA sequencing data used in this manuscript (discovery phase - T1D FinnDiane and DIREVA studies) is provided in the Supplementary Table S3a.

Any additional information required to reanalyze the data reported in this paper is available from the lead contact upon request compatible with the study consents.

## Acknowledgements

We acknowledge the services of University of Helsinki: FIMM Technology Centre HiPREP core for performing EM work and Genomics NGS core for library preparation, sequencing, and primary analysis both supported by HiLIFE and Biocenter Finland, as well as EM Unit of the Institute of Biotechnology for providing the facilities and EV Core for performing the NTA and Exoview. Genevia Technologies Oy is thanked for conducting part of the sequencing data analysis. The authors wish to acknowledge CSC – IT Center for Science, Finland, for computational resources. This project has received funding from the Innovative Medicines Initiative 2 Joint Undertaking under grant agreement No 115974. The JU receives support from the European Union’s Horizon 2020 research and innovation programme and EFPIA and JDRF. Any dissemination of results reflects only the author’s view; the JU is not responsible for any use that may be made of the information it contains. The DIREVA Study (A.K.) has been supported by the Vasa Hospital District, State Research Funding via the Turku University Hospital, Vasa Central Hospital, Jakobstadsnejdens Heart Foundation, the Medical Foundation of Vaasa and the Finnish Medical Foundation. The FinnDiane study has been supported by Folkhälsan Research Foundation, Wilhelm and Else Stockmann Foundation, Liv och Hälsa Society, Sigrid Jusélius Foundation (220027), State funding for university-level health research by Helsinki University Hospital, Novo Nordisk Foundation (NNF23OC0082732), and Diabetes Research Foundation. This study has also been financially supported (TT, OD) by grants from Folkhälsan Research Foundation, the Sigrid Juselius Foundation, The Academy of Finland (grants no. 263401, 267882, 312063, 336822, 312072,336826, 317599), University of Helsinki, Ollqvist Foundation, the Health Care Center in Vasa, and the Finnish Cultural Foundation personal grant (#240316) to KB. This research has been conducted using the UK Biobank Resource under Application Number 170315 (AG). We are grateful for the kind help of Dr Lee, Gachon University College of Medicine, Republic of Korea, and Dr Cortes INCLIVA Biomedical Research Institute, Spain, regarding their published datasets and for the skillful assistance of Jaana Kekkonen, Carola Påhls, Paula Kokko, Anu Dufva, Heli Krigsman, Anna Sandelin, Jaana Tuomikangas, and Kirsi Uljala. We are indebted to the founder of the Direva Study, professor Leif Groop, and the late international coordinator of the FinnDiane Study Group, Carol Forsblom (1964–2022), for their considerable contributions.

